# Pathogenic paralogous variants can be used to apply the ACMG PS1 and PM5 variant interpretation criteria

**DOI:** 10.1101/2023.08.22.23294353

**Authors:** Tobias Brünger, Alina Ivaniuk, Eduardo Pérez-Palma, Ludovica Montanucci, Stacey Cohen, Lacey Smith, Shridhar Parthasarathy, Ingo Helbig, Michael Nothnagel, Patrick May, Dennis Lal

## Abstract

**Purpose:** The majority of missense variants in clinical genetic tests are classified as variants of uncertain significance. Broadening the evidence of the PS1 and PM5 criteria has the potential to increase conclusive variant interpretation.

**Methods:** We hypothesized that incorporation of pathogenic missense variants in conserved residues across paralogous genes can increase the number of variants where ACMG PS1/PM5 criteria can be applied. We mapped over 2.5 million pathogenic and general population variants from ClinVar, HGMD, and gnomAD databases onto 9,990 genes and aligned these by gene families. Subsequently, we developed a novel framework to extend PS1/PM5 by incorporating pathogenic paralogous variants annotations (para-PS1/PM5).

**Results:** We demonstrate that para-PS1/PM5 criteria increase the number of classifiable amino acids 3.6-fold compared to PS1 and PM5. Across all gene families with at least two disease-associated genes, the calculated likelihood ratios suggest moderate evidence for pathogenicity. Moreover, for 36 genes, the extended para-PS1/PM5 criteria reach strong evidence level.

**Conclusion:** We show that single pathogenic paralogous variants incorporation at paralogous protein positions increases the applicability of the PS1 and PM5 criteria, likely leading to a reduction of variants of uncertain significance across many monogenic disorders. Future iterations of the ACMG guidelines may consider para-PS1 and para-PM5.

## Introduction

Large gene panels, exome, and genome sequencing have led to the identification of novel variants at an exponential rate^1^. Up to 80% of pathogenic variants are located within protein-coding regions of the gene^2^, with missense variants being particularly challenging to interpret due to the variety of different molecular mechanisms through which they can cause disease. Furthermore, several disease-associated genes are pleiotropic, which further complicates variant interpretation^3–5^. Despite these challenges, variant classification is necessary for diagnosing rare and genetically heterogeneous disorders, and for the development of personalized medicine.

To standardize variant interpretation, the American College of Medical Genetics and Genomics (ACMG) published recommendations for evaluating the pathogenicity of variants^6^. However, >45% of single nucleotide variants reported in the ClinVar database^7^ (accessed March 2023) are classified as variants of uncertain significance (VUS), due to the absence of sufficient evidence for or against variant pathogenicity. The guidelines include criteria that utilize information from previous genetic test results e.g., the presence of an established pathogenic variant with the same amino acid exchange (PS1) or a different amino acid exchange (PM5) at the same position in the same gene that can provide strong to moderate evidence for pathogenicity^6^. However, since the vast majority of rare monogenic disorders are genetically heterogeneous and about half of the identified pathogenic variants have not yet been observed in other individuals^8,9^, the application of these evidence criteria is limited.

About 80% of genes associated with monogenic disorders are paralogs^10^. These paralogous genes can be grouped into 2871 gene families as defined by the Human Gene Nomenclature Consortium (HGNC)^11^ with >80% sequence similarity^12^. Genes within a gene family arise from gene duplication events of common ancestral genes and can share >90% amino acid sequence similarity at functionally essential protein domains^13^. We and others have demonstrated that functional domains that are conserved within a gene family are enriched for pathogenic variants and depleted for population variants^14–16^. In molecular studies, it has been shown that a protein’s biophysical function at these protein domains is mechanistically conserved within a gene family and therefore, the exchange of a single residue has similar molecular effects across gene family members^16–18^. Therefore, the current PS1/PM5 criteria could be informed not only by prior reported pathogenic variants at the same position in the gene of interest, but also by observed pathogenic variants at conserved positions in related genes of the same family.

Aside from the identification of conservation patterns within gene families, we previously identified differential distribution of missense variants from the general population vs. pathogenic missense variants^16,18^. We showed that evolutionarily conserved protein regions across paralogous genes are enriched for pathogenic variants and depleted for population variants^12^. Further, we identified regions across paralogous genes that are enriched for pathogenic versus control variants, which can be annotated as pathogenic variant enriched regions (PERs)^14^. We demonstrated that new missense variants within PERs are more likely to be pathogenic compared to non-PER regions in the same gene^14^. Although the PER-annotation has the potential to identify disease-sensitive regions across protein sequences, the approach has the disadvantage that it currently has low sensitivity since we were limited in power to identify PERs, as the vast majority of new missense variants are located outside of PERs.

In this new study, we aim to build on previous gene family conservation and PER research to overcome some of the current limitations in clinical variant interpretation. Specifically, we hypothesize that variants in conserved residues in paralogous genes can increase the number of variants classifiable using the ACMG PS1 and PM5 criteria. If our hypothesis is true, the PS1/PM5 ACMG criteria could be extended to include paralogous variants at conserved amino acid positions and increase the number of clinically resolved variants significantly. However, to the best of our knowledge, it has not been tested across the whole protein-coding exome whether single missense variants from paralogous genes can serve as a proxy for variant pathogenicity. As proof of concept, we show that in 519 gene families (1,459 genes) with high sequence similarity, the presence of a single pathogenic variant in a gene family member at a corresponding protein position increases the likelihood that a novel genetic variant at a conserved paralogous position in the target gene is pathogenic.

## Methods

### Gene family definition

We obtained the paralogous genes that belong to a gene family from Pérez-Palma et al. 2020^14^, as originally described in Lal et al., 2020^12^. Briefly, the human paralog definitions were taken from Ensembl BioMart ^19^ and filtered for those with an HGNC symbol^11^. For each gene, the canonical transcript as defined by Ensembl was considered. To avoid aligning highly diverged sequences, families with less than 80% similarity on the full protein sequence were removed.

### Definition of paralogous variants

For all the protein sequences within the same gene family, we performed a multiple sequence alignment using the MUSCLE^20^ software. We then mapped pathogenic and general population variants onto these multiple sequence alignments. Given two variants on two different genes of the same gene family, we considered them as paralogous variants if they satisfied the two following conditions: *1)* they are located at the same position in the multiple protein sequence alignment of the gene family, and *2)* the reference amino acid in the target gene and the paralogous gene is the same (Supplementary Figure 1).

We further establish an expanded set of criteria, termed para-PS1/PM5, which is defined as follows:

**para-PS1:** This refers to a pathogenic paralogous variant that exhibits the same amino acid substitution as the investigated variant.

**para-PM5:** This denotes a pathogenic paralogous variant that exhibits a different amino acid substitution compared to the investigated variant.

### Calculation of the positive likelihood ratio when a pathogenic paralogous variant is found

For each gene, we calculated the positive likelihood ratio using our aggregated set of pathogenic and general population variants for the para-PS1/PM5 criteria (Supplementary Figure 1). While considering the definition of the criteria (see above) we counted for each gene *i)* the number of pathogenic variants (for details about variant collection see Supplementary Methods) for which at least one pathogenic paralogous variant was observed and *ii)* the number of pathogenic variants for which no pathogenic paralogous variant was observed. For the same gene we also counted *i)* the number of control variants for which at least one pathogenic paralogous variant was observed and *ii)* the number of control variants for which no pathogenic paralogous variant was observed. We calculated the positive likelihood ratios following the methodology outlined by Tavtigian et al., 2018^21^ and Pejaver et al.,2022^22^, to determine the level of evidence each criterion provides for pathogenicity. Briefly, we calculated the likelihood ratios in the presence of a pathogenic paralogous variant with either the same amino acid substitution (para-PS1) or a different amino acid substitution (para-PM5). The positive likelihood ratio was computed using the sensitivity and specificity of the test:

Equation 1:

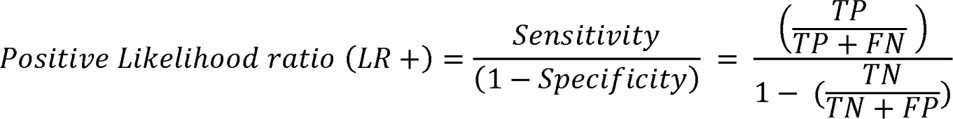

where LR+ represents the positive likelihood ratio, TP denotes true positives, TN signifies true negatives, FP stands for false positives, and FN represents false negatives. To derive a single estimate across all genes, we combined the counts of pathogenic and general population variants and repeated the calculation. All analyses were performed using R v.4.2.1.

## Results

### Incorporating pathogenic paralogous variants increases the number of PS1/PM5 classifiable amino acids

The goal of this study was, through a bioinformatic framework, to test and expand the application of the PS1 and PM5 to paralogous evidence and thus increase the resolution of the current clinical genetic variant interpretation guidelines of the ACMG. Specifically, we investigated whether the presence of pathogenic variants in paralogous genes at a corresponding position can be used to extend the existing strong PS1 (‘same amino acid change as a previously established pathogenic variant regardless of nucleotide change’) and moderate PM5 (novel amino acid substitution where a different amino acid substitution determined to be pathogenic has been seen before) variant pathogenicity classification criteria (Figure 1). We defined a ‘paralogous variant’, as a variant that fulfills two criteria: (1) it is located in a paralogous gene at the corresponding residue index position, as determined by multiple sequence alignment (see Methods for details), and (2) with the same reference amino acid as in the target gene.

**Figure 1:**
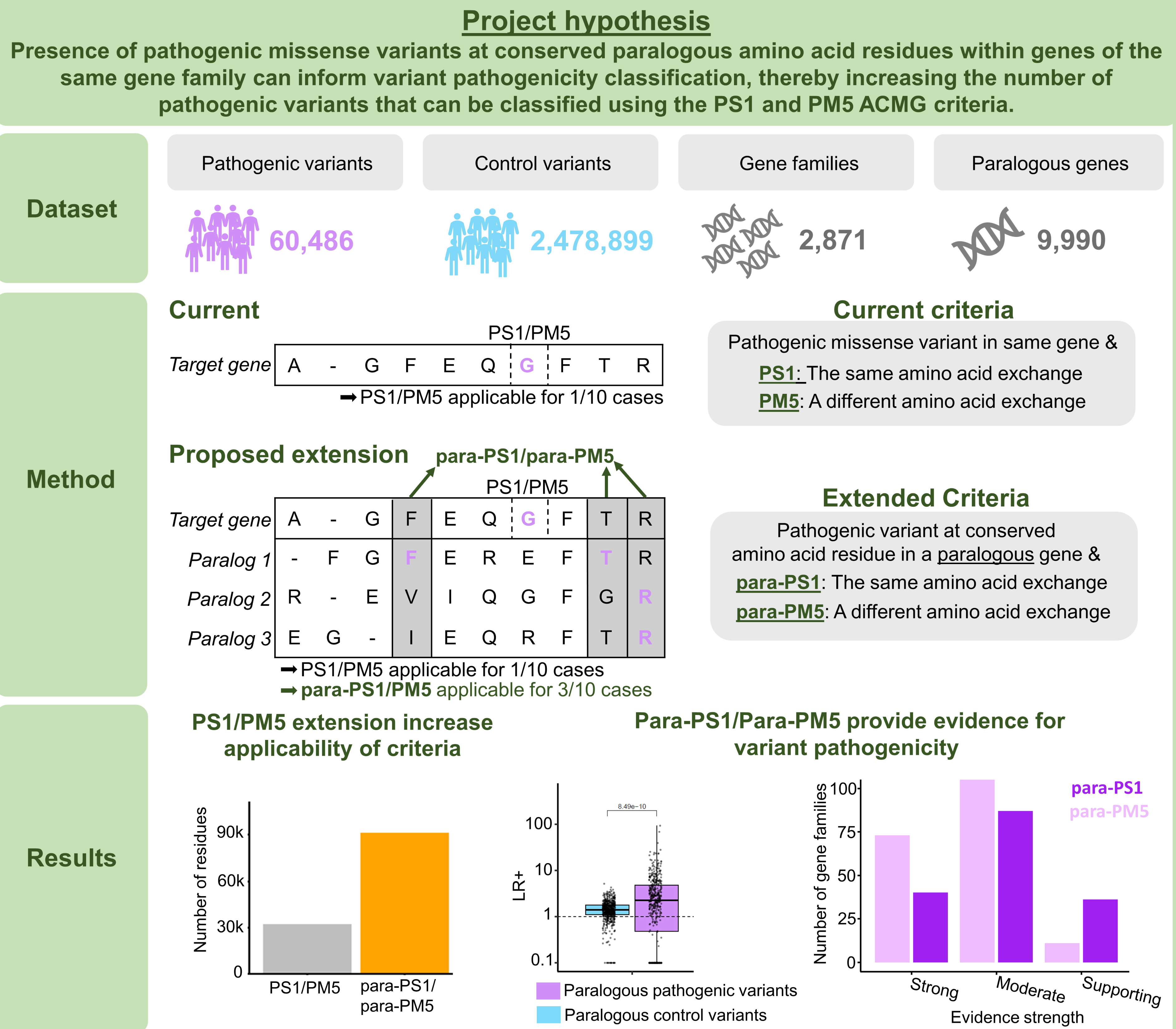
**Graphical summary of the study.**

The inclusion of paralogous variants could decrease the number of variants of uncertain significance. Before establishing the validity of the incorporation of paralogous variants in the PS1/PM5 criteria, we quantified the number of additional amino acids residues where the updated PS1/PM5, further referred to as para-PS1 & para-PM5, could be applied. We aggregated a total of 60,486 pathogenic variants from ClinVar^7^ and HGMD^23^ and mapped them to 2,871 different gene family alignments, consisting of 9,990 genes (Figure 1). Our paralog variant analysis integrates pathogenic variants from multiple genes in the same gene family (see Methods for details). We, therefore, restricted the dataset to gene families harboring pathogenic variants in at least two genes and identified 1,459 genes from 519 gene families. Within these genes, 41,223 pathogenic missense variants and 171,690 pathogenic paralogous variants were found that covered 32,137 and 91,259 amino acid residues respectively (Supplementary Table 1). Of these 91,259 residues that are covered by a paralogous pathogenic variant 92.6% (N = 84,553 residues) were not covered by a pathogenic variant in the same gene. Therefore, the application of the extended ‘para-PS1’ and ‘para-PM5’ criteria would increase the number of amino acids in these gene families were the criteria can be applied by about 3.6-fold (N = 116,690 residues, Figure 2A). The increase in the number of classifiable amino acids in each gene family is highly correlated with the number of disease-associated genes in a gene family (R = 0.97, P = <1e-300, Supplementary Figure 2).

**Figure 2:**
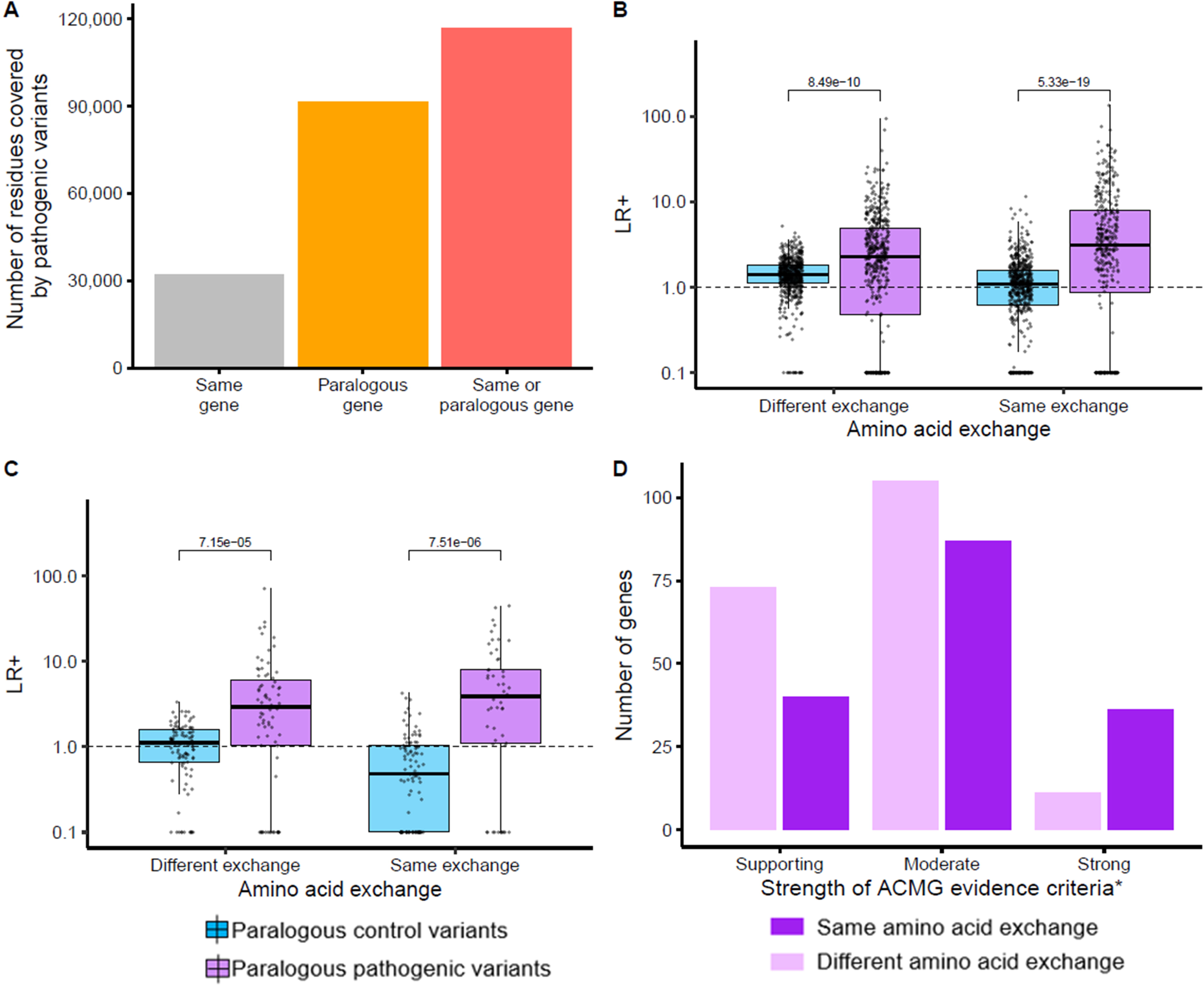
Individual pathogenic paralogous variants can serve as a proxy for variant pathogenicity. A) Number of amino acid residues in 519 gene families that have a pathogenic variant (ClinVar/HGMD) at the same protein position in the same gene or a corresponding protein residue in a paralogous gene. B) Amino acids with a paralogous pathogenic variant at a paralogous aliment position have an increased positive likelihood ratio (LR+ >1). In contrast, amino acids with a paralogous control variant (gnomAD) at a paralogous alignment position are not enriched for pathogenic variants. Each data point represents the gene-wise LR+. The gene-wise LR+ was calculated for genes where 10 or more pathogenic variants (ClinVar/HGMD) and control variants (gnomAD) could be mapped. C) As in (B), but limited to missense constraint genes (Missense-z score > 3.09). D) Barplot showing the number of genes where pathogenic variants with the same (Para-PS1, purple) or different amino acid exchange (para-PM5 pink) provide supporting-strong evidence for variant pathogenicity. *Following the Bayesian framework proposed by Tavtigian et al., 2018.

### Presence of a single pathogenic paralogous variant can provide supportive to strong evidence for variant pathogenicity

Next, we tested the validity of the inclusion of pathogenic paralogous variants for the application of the para-PS1 & para-PM5 criteria. In addition to the aforementioned pathogenic variants, we included 2,478,899 general population variants from the gnomAD database^24^, that served as controls in our study. To estimate the evidence of the para-PS1 and para-PM5 criteria we positive likelihood ratio (LR+) following the Bayesian framework proposed by Tavtigian et al. in 2018^21^. Across the 519 selected gene families, we observed an LR+ of 8.32 (8.02-8.62, 95% confidence interval (CI)) pathogenic versus control variants when a pathogenic paralogous variant with the same amino acid exchange was present at a corresponding alignment index position (para-PS1, for details on the approach, see Methods). Restricting the analysis to missense variant-constrained genes (Missense-z score > 3.09^1^), increased the LR+ to 8.91 (8.03-9.88, 95% CI). Applying proposed cutoffs to translate the LR+ to weights of evidence considering the suggested^21^ prior probability of 0.1, we can consider an LR+ > 18.7 as strong evidence, LR+ > 4.3 as moderate, and LR+ > 2.08 as supporting evidence for variant pathogenicity^21^. Consequently, across all assessed genes the para-PS1 criterium can be considered as moderate evidence. Notably, even for paralogous variants with a different substitution at the same alignment index position (para-PM5), we observed an increased LR+ (All genes: LR+ = 4.32, (4.24-4.48, 95% CI), Missense constraint genes: LR+ = 6.48, (6.05-6.94, 95% CI). The para-PM5 criterium could also be considered as moderate evidence if the point estimate is considered and as supporting evidence if the lower bound of the 95% interval (LR+_lower bound_ = 4.24) is considered.

Next, we calculated for a subset of genes for which at least 10 pathogenic and control variants could be mapped on the gene LR+ on a gene-wise level. We observed a wide range of LR+ across different genes (Figure 2A, B). Pathogenic paralogous variants applied as para-PS1 criterium (same amino acid exchange), provide strong evidence for variant pathogenicity in 36 genes, moderate evidence in 87 genes, and supporting evidence in 40 genes (Figure 2D, Supplementary Table 2). After assessing the same amino acid exchange, we moved on to evaluating the evidence for pathogenic paralogous variants at alignment index positions with different substitutions, applying the para-PM5 criteria. We observed that while the para-PS1 same substitution criteria provided strong evidence in 24.5% of genes with an LR+ >2.08 (N = 163 genes), whereas the para-PM5 criteria provided strong evidence in only in 5.8 % of genes (N genes _with para-PM5 LR+ >2.08_ = 187 genes). Overall, genes where paralogous pathogenic variants can provide strong evidence under either para-PS1 and/or para-PM5 criteria typically contained fewer genes and also tended to contain fewer genes where pathogenic variants are established (Supplementary Figure 3 A-D). Notably, genes with higher constraints for missense variants did not show significantly higher positive likelihood ratios (Supplementary Figure 3 E, F).

### The para-PS1/para-PM5 improves previous family-based variant interpretation approaches

Previously, we developed two methods for variant classification, both of which harnessed the power of gene family information on an exome-wide scale^12,14^. Initially, we established a paralog conservation score (referred to as ‘Parazscore^12^’) and demonstrated that amino acids conserved within a gene family are significantly enriched for pathogenic variants. Notably, a fundamental prerequisite for the implementation of para-PS1 & para-PM5 criteria is the conservation of amino acid residues between the target gene and its paralogous gene, in which a paralogous pathogenic variant is detected. Hence, whenever para-PS1 or para-PM5 criteria become applicable, a certain degree of conservation within the genes of the same gene family becomes inevitable. This conservation potentially explains a portion of the elevated LR+ we observed. To further elucidate the potential impact of paralog conservation on the calculated LR+, we repeated our previous analysis allocating every amino acid to subgroups depending on their paralog conservation. This segmentation was done using the Parazscore as a metric to quantify paralog conservation (see Supplementary Methods for details). Interestingly, within these subgroups, the highest LR+ were observed for residues exhibiting the least paralog conservation for both the para-PS1 criterium (Parazscore <0; LR+_para-PS1_ = 10.49, 95% CI = 9.60-11.45, Figure 3A) as well as the para-PM5 criterium (Parazscore <0; LR+_para-PM5_ = 5.21, 95% CI = 4.87-5.59, Figure 3B). Yet, even within the subgroup demonstrating the least increase in LR+, where maximum conservation across all paralogous genes of the same gene family was noted, we still detected an increased LR+ of 4.88 and 2.69, for para-PS1 and para-PM5 respectively. This observation suggests that the existence of pathogenic paralogous variants provides additional information beyond the level of conservation between paralogous genes.

**Figure 3:**
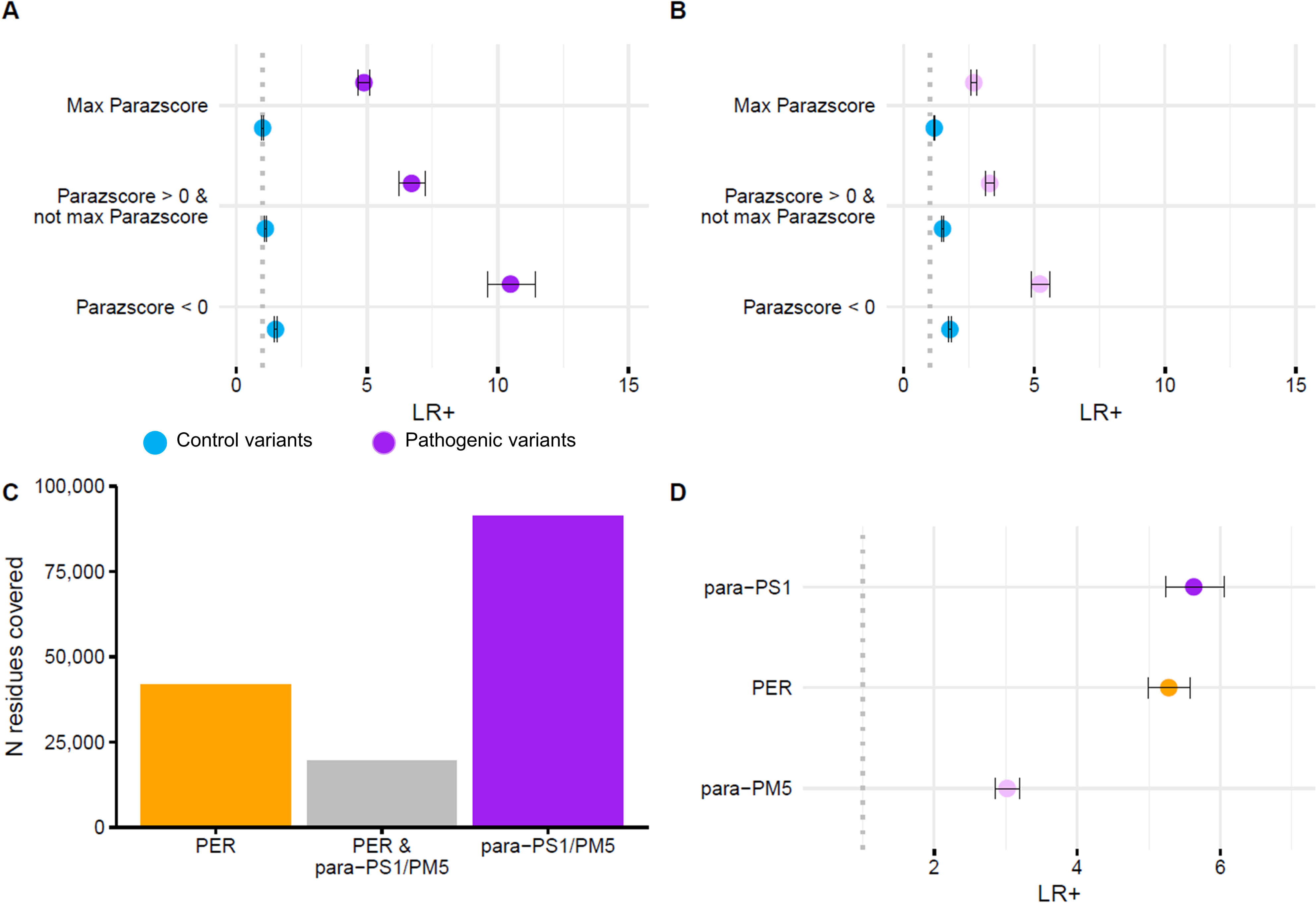
Comparison to established gene family-based methods. A) The forest plot illustrates the enrichment of pathogenic versus control variants applying para-PS1 for residues with similar paralog conservation levels, as defined in Lal et al., 2020^12^. B) Similar to (A), but for the para-PM5 criterium. C) The bar plot shows the number (N) of amino acid residues across all genes where a previously established approach (Pathogenic Enriched Region, PER; Perez-Palma et al., 2019^14^) and/or our para-PS1/ para-PM5 ACMG criteria extension can be applied. D) The forest plot compares the likelihood ratios (LR+) for amino acid residues within a PER and amino acid residues where para-PS1/para-PM5 criteria can be applied (see Supplementary Methods for details).

We then compared the para-PS1/PM5 criteria with a second approach^14^ which utilizes pathogenic variants from paralogous genes of the same gene family. By analyzing the distribution of pathogenic and control variants across a gene family the approach identifies pathogenic variant enriched regions (‘PERs’, on average 33 consecutive amino acids^14^) across paralogous genes that are significantly enriched for pathogenic versus control variants. Due to the sliding window approach to identifying regional enrichments of pathogenic variants, PERs can span amino acid residues without an established pathogenic variant across paralogs, and the regional association is derived from adjacent variants. However, identifying PERs within a gene family alignment requires a large number of pathogenic variants, limiting its applicability. First, we compared the number of exome-wide classifiable variants using para-PS1/PM5 with the PER approach. We used an independent set of pathogenic and control variants that were not utilized in the PER generation or the generation or validation of the para-PS1 and para-PM5 criteria (see Supplementary Methods for details). We found that the para-PS1 & para-PM5 approaches captured 2.2 times more residues compared to PERs (Figure 3C). In the second comparison, we compared the LR+ for each approach and observed similar LR+ for the PER approach and for the para-PS1 approach, both suggesting moderate evidence for pathogenicity (LR+_PER_ = 5.28, LR+_para-PS1_ = 5.63, Figure 3D). In contrast, para-PM5 with an LR+ of 3.02 could provide supporting evidence for pathogenicity.

### Application of para-PS1/para-PM5: Two case examples

To demonstrate how the ACMG criteria extension para-PS1 & para-PM5 could lead to a change in variant classification, we provide two examples where our para-PS1 and para-PM5 have been applied (Figure 4).

**Figure 4:**
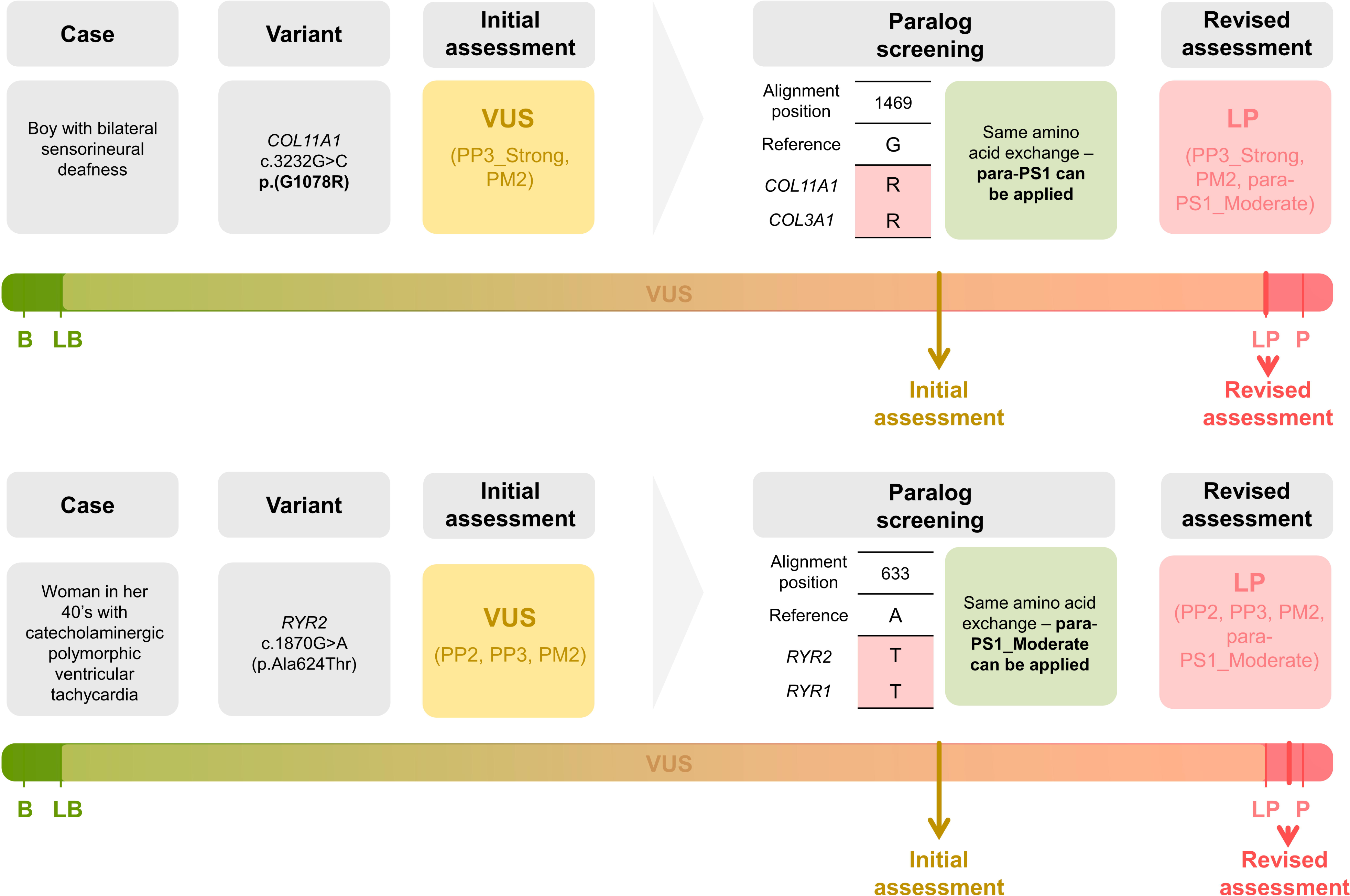
Case examples to demonstrate the utility of the para-PS1/PM5 criteria. Upper panel: Case 1, a boy with bilateral sensory hearing loss and a heterozygous variant in *COL11A1* that is initially classified as a variant of uncertain significance (VUS). By considering a pathogenic paralogous variant found in *COL3A1 and COL1A2* and consequently applying the para-PS1 criterium (moderate evidence) the variant could be reclassified to likely-pathogenic (more details in the results section). Lower panel: Case 2, a women in her 40’s with syncopal episodes, a recent diagnosis of catecholaminergic polymorphic ventricular tachycardia and a heterozygous variant in RYR2 gene, NM_001035.3:c.1870G>A (p.Ala624Thr) that is initially classified as VUS. By considering a pathogenic paralogous variant with the same amino acid exchange found in RYR1 we could apply the para-PS1 criterium and reclassify the variant to likely-pathogenic. Abbreviations: B = benign, LB = likely-benign, VUS = variant of uncertain significance, LP = likely-pathogenic, P = Pathogenic.

Case 1 is a young boy assessed for bilateral sensory hearing loss with a presumably infantile onset. Genetic testing revealed a heterozygous variant in *COL11A1* NM_001854.4:c.3232G>C (p.Gly1078Arg). The variant is absent in the gnomAD database^24^ (moderate criterion PM2) and multiple lines of computational evidence support its pathogenicity (REVEL^25^= 0.997, MutPred2^26^= 0.975 – strong evidence PP3_Strong^22^). The available evidence (Supplementary Table 3) would allow a classification of the variant as a variant of uncertain significance (VUS) and leave the diagnosis uncertain^6^. However, several paralog genes have pathogenic variants reported at the same alignment position (1469) (Supplementary Table 1). Variants in two paralog genes have the same exchange: *COL3A1* (p.Gly729Arg) and *COL1A2* (p.Gly643Arg). According to our findings (Supplementary Table 2), this can be applied as a criterion para-PS1_Moderate, which would allow us to classify the variant as likely pathogenic scoring 2 moderate (para-PS1_Moderate, PM2) and one strong (PP3_Strong) criteria.

Case 2 is a woman in her 40’s with syncopal episodes since childhood and a recent diagnosis of catecholaminergic polymorphic ventricular tachycardia. She underwent a genetic panel testing revealing a heterozygous variant in *RYR2*, NM_001035.3:c.1870G>A (p.Ala624Thr). No other direct family members were available for testing. *In-silico* prediction tools provide supporting evidence for variant pathogenicity (REVEL^25^= 0.573, MutPred2^26^= 0.799; PP3 ^22^). The variant affects a missense-constraint gene^27^ (PP2) and is not present in gnomAD^24^ database (PM2). Taken together, available evidence (Supplementary Table 3) classifies the variant as VUS However, we observed a pathogenic variants with the same exchange in the paralogous gene *RYR1* p.Ala612Thr (Supplementary Table 1), which allows us to apply a para-PS1_Moderate (Supplementary Table 2). Considering the application of the criterion, the variant can be classified as likely pathogenic given 2 supporting (PP2, PP3) and 2 moderate (PM2, para-PS1_Moderate) criteria.

## Discussion

Many paralogs are highly conserved in sequence and have similar biophysical molecular functions. However, the current variant interpretation guidelines consider only pathogenic missense variants in the gene of interest. Here, we developed and validated a bioinformatic framework to include missense variants from paralogous genes at alignment index positions as additional source for the application of the PS1 and PM5 criteria (‘para-PS1’ and ‘para-PM5’). Our analysis revealed that the extended para-PS1 (pathogenic paralogous variant with the same amino acid exchange as the target variant) and para-PM5 (pathogenic paralogous variant with a different amino acid exchange as the target variant) criteria can increase the number of residues where the criteria can be applied compared to original PS1 and PM5 criteria by 3.6 fold. We show that the para-PS1 and para-PM5 criteria can provide supporting to strong evidence of pathogenicity and complement and extend existing methods^12,14^.

Despite efforts to standardize criteria for pathogenicity assignment^6^ and many improvements in variant interpretation, about 75% of missense variants in ClinVar^7^ (accessed 12/2022) are annotated as variants of uncertain significance (VUS). Extending or modifying existing ACMG criteria has been demonstrated as a promising approach to reclassifying VUSs ^22,28–31^. We demonstrated that the PS1 and PM5 criteria of the ACMG guidelines, which consider knowledge about an established pathogenic variant with the same amino acid change as a strong evidence criterion for variant pathogenicity, can be extended by including paralogous pathogenic variants.

According to the current guidelines^6^, the presence of a pathogenic variant with the same amino acid substitution in the same gene (PS1) provides strong evidence of pathogenicity, whereas the presence of a pathogenic variant with a different amino acid substitution in the same gene (PM5) provides moderate evidence^6^. This is consistent with our observation that the para-PS1 enrichment for pathogenic variants was higher (OP = 8.32) compared to the enrichment that we observed when the para-PM5 criterion was applied (OP = 4.32). To estimate the level of evidence the extended para-PS1 criteria could provide we followed the Bayesian framework as proposed by Tavtigian et al., 2018^21^ and previously applied in several other studies^32–35^. In contrast to the established PS1 and PM5 criteria, which provide strong and moderate evidence respectively, both para-PS1 and para-PM5 can provide moderate evidence across genes in our study. We speculate that the difference between the moderate evidence provided by para-PS1 criterium compared to the strong evidence by PS1 may be due to potentially misclassified variants in large patient variant databases such as ClinVar^36^ which affects the quantification of the reported effect. A problem was observed for novel variants submitted before the availability of large reference databases. Importantly, while the level of evidence for variant pathogenicity that PS1 and PM5 can provide is considered uniform across all genes^6^ we showed that for the extended criteria, the level can be calculated specifically for each gene family. For example, while the positive likelihood ratio across all gene families meets the evidence threshold for moderate strength, for 36 genes the LR+ is >18.7, which can be translated into strong evidence^21^.

Pathogenic missense variants in paralogous genes can serve as a proxy for pathogenicity. Within a protein sequence, pathogenic variants are unevenly distributed and tend to accumulate in certain regions that are critical for protein function^37^. These pathogenic variant-enriched regions have proven valuable for variant classification through established guidelines for variant interpretation^6^ and the use of *in-silico* prediction algorithms^38^. Moreover, the observation that critical protein regions tend to be evolutionarily conserved between paralogous genes can be harnessed to enhance statistical robustness by incorporating pathogenic variants across these paralogous genes^14^. Still, about 70%, of pathogenic variants are located outside the regions identified as essential. As a result, individual pathogenic variants in paralogous genes outside these regions were not considered for variant interpretation. In a study examining long QT syndrome, it was observed that individual pathogenic variants in paralogous genes are often located at paralogous positions as determined from multiple sequence alignments^16^, suggesting that the presence of a pathogenic variant at a particular position may serve as a proxy of pathogenicity at that alignment position in other paralogs. Our data test this hypothesis across a wide range of gene families and suggests that individual pathogenic paralogous variants can indeed serve as proxies for pathogenicity on a broad scale, thereby augmenting the efficacy of established variants in variant interpretation frameworks.

Our proposed inclusion of the paralogous variants as an extension of the PS1 and PM5 ACMG criteria in future iterations comes with a few limitations. One limitation is the focus on paralog conserved regions. Our approach does not capture pathogenic variants in gene-specific regions that are not conserved across paralogous genes. Within these regions, the original PS1/PM5 criteria can be applied whereas the extended para-PS1/PM5 criteria can not. Our approach is an extension of the PS1/PM5 criteria and does not limit their application in such regions. Furthermore, our gene families were required to have >80% sequence similarity and previous studies have shown that the majority of functionally important regions are conserved across paralogs^12^, and *de novo* domains are rare and could be annotated separately. Secondly, a limitation of our study is the inclusion of control variants aggregated from the gnomAD database, some of which may be pathogenic despite their presence in the general population. In instances where these control variants are indeed pathogenic, the likelihood ratios calculated in our study may represent underestimations, maintaining the conservative nature of our findings. Another limitation is the partially small number of established pathogenic variants and paralogous pathogenic variants. Many gene families lack a sufficiently large collection of identified pathogenic variants that serve as a proxy for pathogenicity which limits statistical power to confidently quantify the gene-specific evidence provided by paralogous pathogenic variants. The number of individual genes where paralogous pathogenic variants provide significant supportive or higher evidence for pathogenicity was associated with the number of paralogous pathogenic variants (P_para-PS1_= 1.6e-24, P_para-PM5_= 4.6e-09, Supplementary Figure 4). Consequently, although we have demonstrated that our approach can be powerful to extend the current ACMG criteria, there are individual genes where our approach lacks sufficient power to validate evidence strength on the gene level, as indicated by 169 genes that have increased LR+ when a paralogous pathogenic variant with the same exchange is present, however without statistical significance.

In conclusion, our findings suggest that the extended criteria para-PS1 and para-PM5 have significant potential to improve variant interpretation and aid in the diagnosis of pathogenic variants in clinical practice. Reference databases continue to grow and include well-classified pathogenic variants. While the applicability of the PS1 and PM5 criteria will, as a consequence, will grow the applicability of the para-PS1/PM5 criteria will increase at an even faster rate. Future iterations of variant interpretation guidelines that consider the presence of paralogous pathogenic variants as evidence of pathogenicity could thus significantly increase the application of criteria based on already established pathogenic variants.

## Declaration of generative AI and AI-assisted technologies in the writing process

During the preparation of this work, the authors used CHAT-GTP-4 to improve readability and language. After using this tool/service, the authors reviewed and edited the content as needed and take full responsibility for the content of the publication.

## Data availability

Data is available in the Supplementary Tables.

## Author Contributions

Conceptualization: T.B., A.I., D.L. I.H.; Data curation: T.B., A.I., E.P; Analysis: T.B, A.I., Supervision: D.L.; P.M.; M.N.; Writing-original draft: T.B., A.I., L.M.; Writing-editing: D.L., P.M., S.C, L.S., S.P., L.M.

## Ethics Declaration

This study was approved by Cleveland Clinic IRB, approval ID 22-147. Informed consent was waived considering the retrospective nature of the study.

## Confict of Interest

The authors report no conflicts of interests.

