## Supplementary Material for "Pathogenic paralogous variants can be used to apply the ACMG PS1 and PM5 variant interpretation criteria"

### Supplementary Methods

#### Annotation of missense variants from public repositories

##### *Missense variants from patients*

Missense variants associated with the disease were collected from the ClinVar database[7] (ClinVar, release October 2019) and the Human Gene Mutation Database[19] (HGMD®) Professional release 2019.2. Similarly, we gathered an updated version of the variants from ClinVar (released December 2022) and HGMD (Professional release 2023.1), processed them as described before, and extracted all previously unreported pathogenic variants not observed in the previous dataset to obtain an independent set of variants. The ClinVar missense variants were obtained in a tabular format from the FTP site (ftp://ftp.ncbi.nlm.nih.gov/pub/clinvar/) and only those variants exclusively classified as "Pathogenic" and/or "Likely Pathogenic" in their final consensus interpretation were considered to ensure high stringency. The HGMD dataset was filtered for "missense variants," "High Confidence" calls (hgmd_confidence = "HIGH" flag), and "Disease causing" state (hgmd_variantType = "DM" flag). All annotations refer to the human reference genome version GRCh37.p13/hg19, and variants belonging to non canonical transcripts as defined by Ensembl were excluded[20]. Since ClinVar and HGMD are not mutually exclusive, we used the union of both resources and removed duplicate entries by comparing HGVS annotations. We further refer to the combined set of variants classified as likely-pathogenic, pathogenic, or "Disease-causing" as ‘pathogenic variants’.

##### *Missense variants from the population*

Missense variants present in the Genome Aggregation Database[21] (gnomAD, public release 2.0.2) were obtained in the Variant Call Format[22] (VCFs). We extracted the high-quality missense variants by filtering the VCF files to the "CSQ" field and “PASS” flag. The annotations were based on the human reference genome version GRCh37.p13/hg19. We extracted only entries annotated to the canonical gene transcripts, as defined by Ensembl[20]. The aggregated population variants serve as control variants in our study and are further referred to as controls.

Similarly, we gathered an updated version of the variants from gnomAD (public release 2.1.1, processed them as described above, and extracted all novel variants not observed in the previous set of gnomAD variants to obtain an independent set of control variants.

#### Comparison to established gene-family-based approaches

To compare our results to an established gene-family-based approach which identified pathogenic enriched regions (PERs) across paralogous genes on an exome-wide scale[14], we gathered an independent set of variants (see Annotation of missense variants from public repositories) which was not previously used nor in the PER approach nor the enrichment analysis of this study, and we repeated the calculation outlined above.

To estimate LR+ that are not mediated by paralog conservation we repeated the analysis described above for three paralog conservation sub-groups using the Parazscore[12]. The groups we considered are alignment positions with gene family wise 1) maximum Parazscore, indicating full paralog conservation across the gene family at the alignment position 2) Parazscore>0 & not maximum Parazscore, indicating high paralog conservation at this alignment position but not full conservation and 3) Parazscore<0, indicating low levels of conservation between paralogous genes at the alignment position.

### Collection and classification of case examples

Two cases to illustrate the usage of paralog-based PM5 and PS1 criteria were selected among the individuals who received care in the Cleveland Clinic healthcare network (CCF) and were found to have VUS in genes related to their disorder in the years 2018-2023. We chose to focus on genes related to cardiac arrhythmias and collagenopathies considering the clinical focus of the CCF centers and the corresponding high patient volume. The variants were extracted and re-analyzed by a clinician trained in clinical variant interpretation (A.I.). ACMG/AMP Standards and Guidelines for the Interpretation of Sequence Variants were used for classification; any used specifications and modifications for the usage of particular criteria are cited as appropriate. The clinical data used for assessment was de-identified and limited to the minimum required for variant classification. The data was stored and processed using secure in-house infrastructure This part of the study was approved by the Cleveland Clinic IRB, approval ID 22-147. Informed consent was waived considering the retrospective nature of the study.

### Supplementary Tables legends

**Supplementary Table 1: List of amino acid residues overlapping with paralogous (likely)-pathogenic variants from ClinVar or HGMD databases.** For each residue the table displays all conserved missense variants in paralogous genes that have been previously annotated as (likely)-pathogenic in the ClinVar or HGMD database. If for a target residue multiple variant are found they are separated by a semicolon.

**Supplementary Table 2: Evaluation of evidence strength for para-PS1/PM5 criteria on a gene-wise basis.** This table presents the evidence strength for the para-PS1/PM5 criteria, focusing on those genes where these criteria provide at least significant supporting evidence for pathogenicity at the gene level. *The methodology employed for this evaluation follows the approach proposed by Tavtigian et al., 2018.

**Supplementary Table 3: Initial assessment of clinical significance of variants included in the case examples.**

### Supplementary Tables

**Supplementary Table 3: Initial assessment of clinical significance of variants included in the case examples.**

| | **Case 1** | **Case 2** |
|---|---|---|
| Gender | Male | Female |
| Clinical features | Bilateral sensorineural hearing loss, mild developmental delay | Catecholaminergic polymorphic ventricular tachycardia |
| Detection method | Gene panel | Gene panel |
| cDNA and protein change | *COL11A1* NM_001854.4:c.3232G>C (p.Gly1078Arg) | *RYR2* NM_001035.3:c.1870G>A (p.Ala624Thr) |
| Inheritance | Unknown | Unknown |
| ACMG Evaluation | VUS | VUS |
| Applied criteria | PM2, PP3_Strong | PM2, PP3, PP2 |

### Supplementary Figures

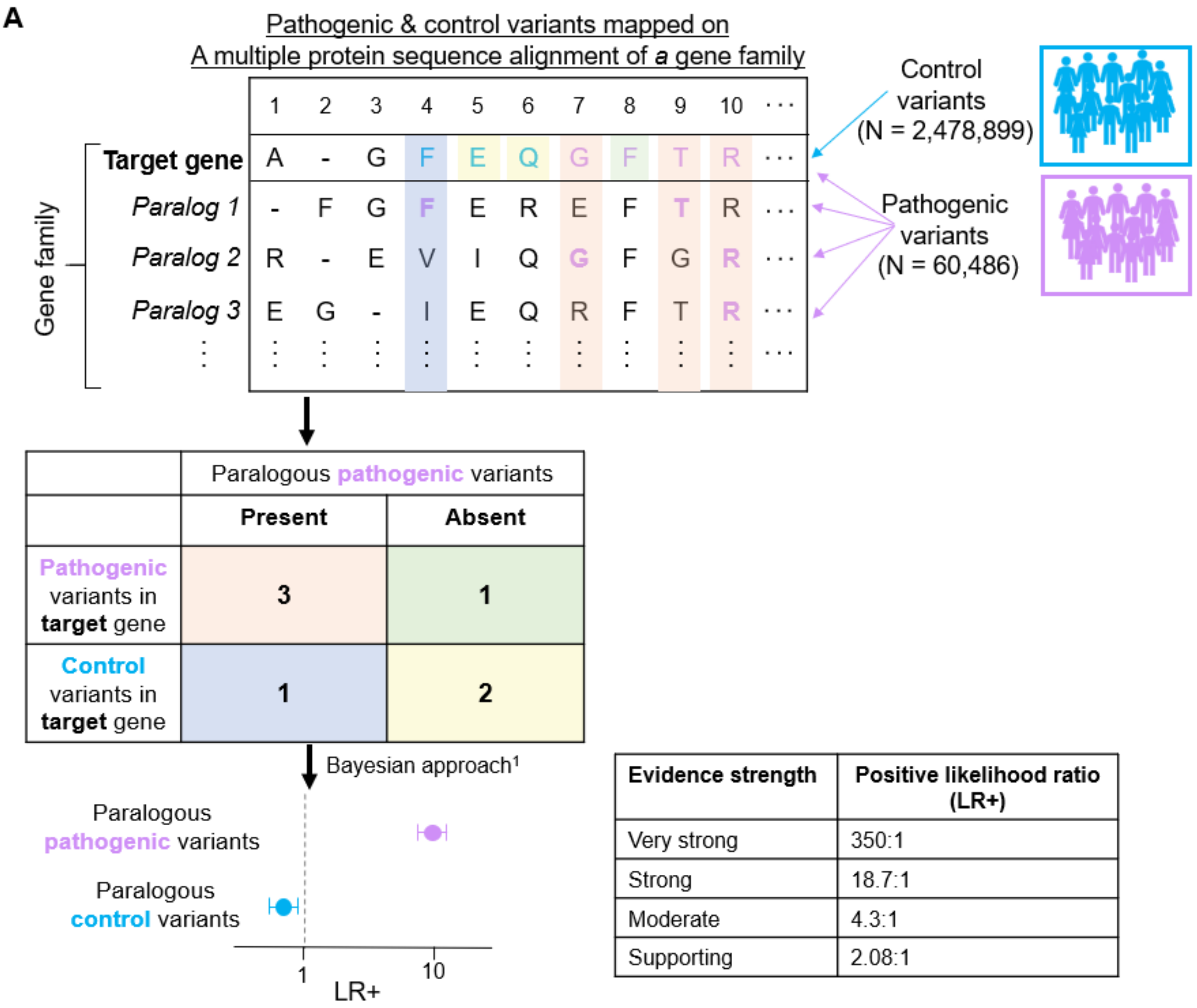


| Evidence strength | Positive likelihood ratio (LR+) |
|---|---|
| Very strong | 350:1 |
| Strong | 18.7:1 |
| Moderate | 4.3:1 |
| Supporting | 2.08:1 |

**Supplementary Figure 1: Calculation of the likelihood ratio in the presence of pathogenic paralogous variants.** Evidence strengths were obtained from Tavtigian et al., 2018 and considered a prior probability of 0.1. 1Approach applied to determine the strength of ACMG criteria in Tavtigian et al. 2018.

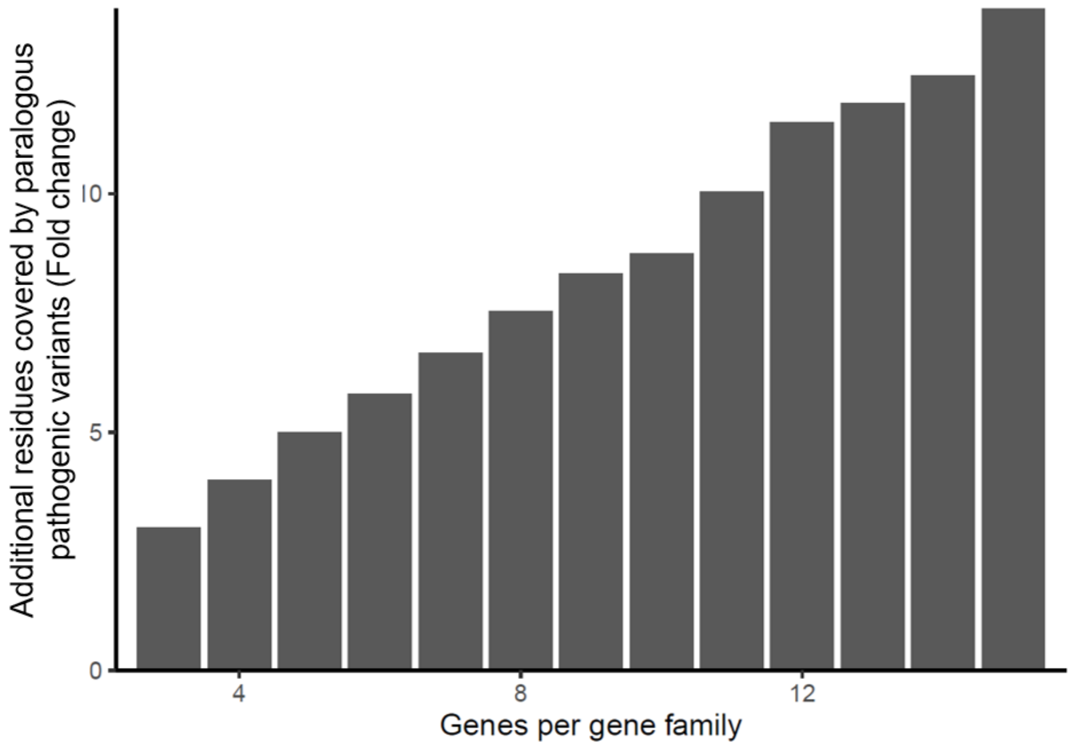


**Supplementary Figure 2: The barplot shows a fold-wise increase in pathogenic variant residue coverage with consideration of paralog genes compared to the same gene only.** The fold-change is stratified by the number of genes in a gene family.

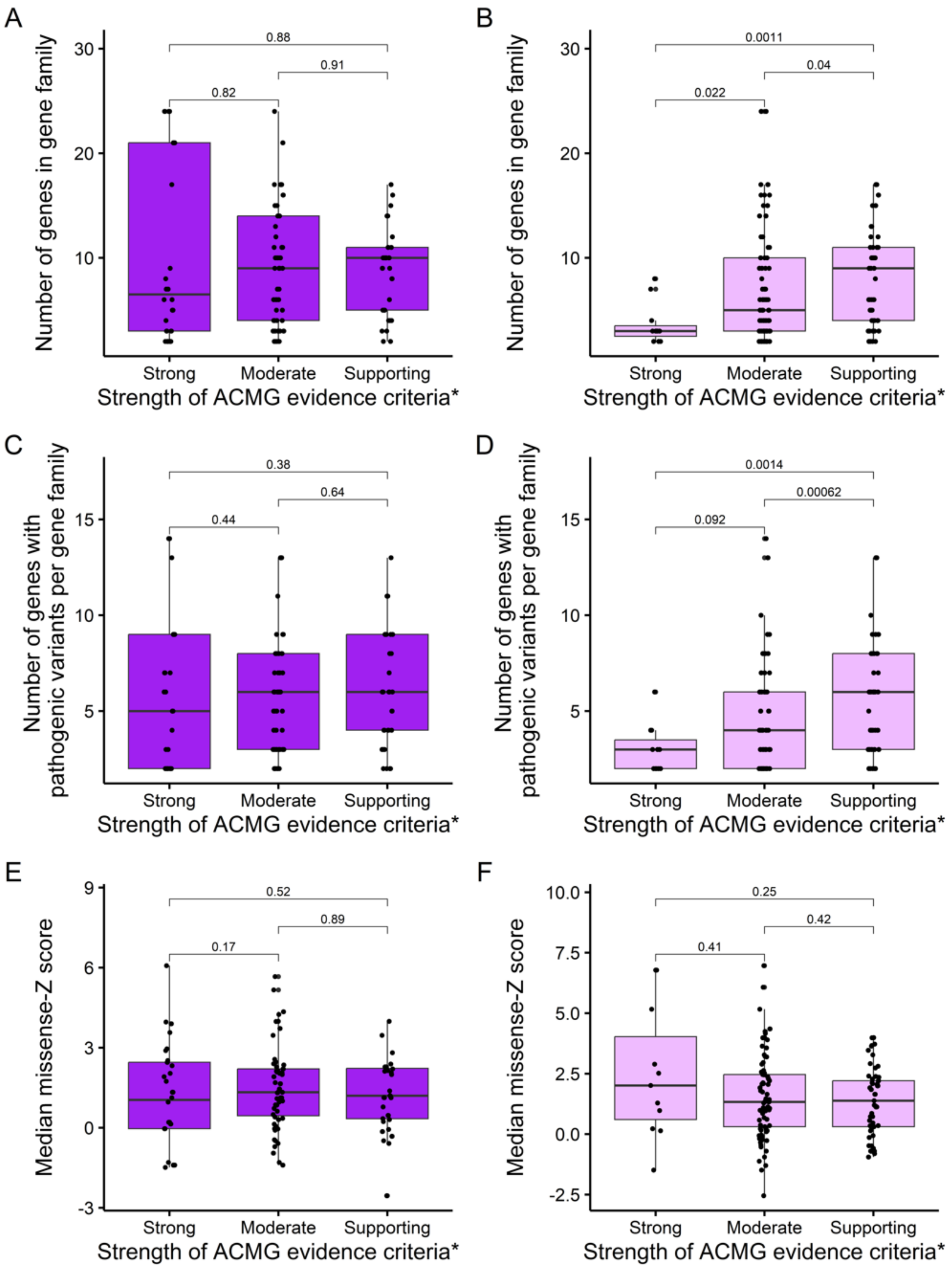


**Supplementary Figure 3: Variability in the strength of the ACMG evidence criteria (para-PS1/para-PM5) across different genes.** A) This panel displays a boxplot representation illustrating the relationship between the number of genes within each gene family and the strength of pathogenicity evidence as calculated for the para-PS1 criteria (same amino acid substitution). B) Similar to Panel A, but demonstrates the association for para-PM5 criteria (different amino acid substitution). C) A boxplot showcasing the correlation between the quantity of observed pathogenic variants per gene and the strength of pathogenicity. evidence based on the para-PS1 criteria. D) Analogous to Panel C, but for para-PM5 criteria. E) A boxplot representing the median gene-level missense constraint (missense-Z score) in relation to the strength of para-PS1 criteria. F) Mirroring Panel E, but in the context of para-PM5 criteria.

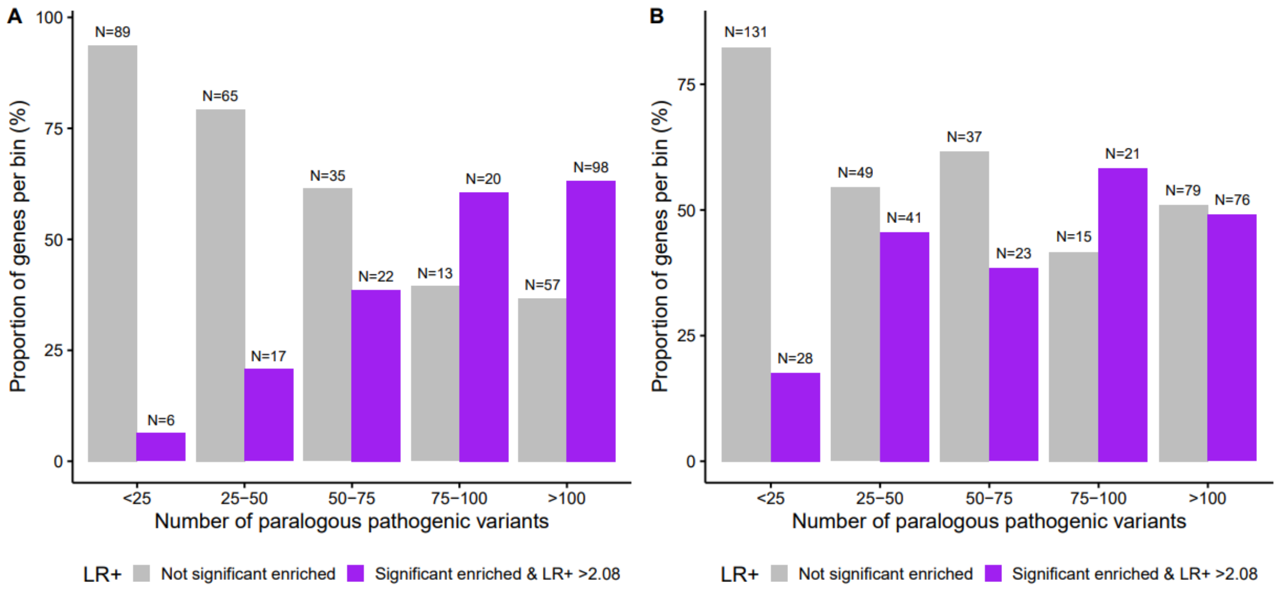


**Supplementary Figure 4: Number of genes where para-PS1/PM5 provide statistically significant supporting evidence correlates with the number of paralogous pathogenic variants.** The barplot shows the proportion of genes per group that have a significantly increased positive likelihood ratio (LR+) (LR+ >2.08 and LR+ lower 95% confidence interval >1). Genes that provide at least significant supporting evidence are shown in purple. Genes that do not are colored in grey.
